# Association between common adverse events after COVID-19 vaccination and anti-SARS-CoV-2 antibody concentrations in a population-based prospective cohort study in the Netherlands

**DOI:** 10.1101/2023.10.19.23297194

**Authors:** M.R. Holwerda, C.E. Hoeve, A.J. Huiberts, G. den Hartog, H.E. de Melker, S. Van den Hof, M.J. Knol

## Abstract

**INTRODUCTION:** Adverse events (AE) such as pain at injection site or fever are common after COVID-19 vaccination. We aimed to describe determinants of AE after COVID-19 vaccination and investigate the association between AE and pre- and post-vaccination antibody concentrations.

**METHODS:** Participants of an ongoing prospective cohort study (VASCO) completed a questionnaire on AE within two months after COVID-19 vaccination and provided 6-monthly serum samples. Data from May 2021 to November 2022 were included. Logistic regression analyses were performed to investigate determinants of AE after mRNA vaccination, including pre-vaccination Ig antibody concentrations against the receptor binding domain. Multivariable linear regression was performed in SARS-CoV-2 naïve participants to assess the association between AE and log-transformed antibody concentrations 3-8 weeks after mRNA vaccination.

**RESULTS:** 47,947 AE questionnaires were completed by 28,032 participants. In 42% and 34% of questionnaires, injection site and systemic AE were reported, respectively. In 2.2% of questionnaires, participants sought medical attention due to AE. AE were reported significantly more frequently by women, younger participants (<60 years), participants with medical risk conditions and Spikevax recipients (versus Comirnaty). Higher pre-vaccination antibody concentrations were associated with higher incidence of systemic AE after the second and third dose, but not with injection site AE or AE for which medical attention was sought. Any AE after the third dose was associated with higher post-vaccination antibody concentrations (geometric mean concentration ratio: 1.38, 95%CI 1.23-1.54).

**CONCLUSION:** Our study suggests that high pre-vaccination antibody levels are associated with AE, and that experiencing AE may be a marker for a good antibody response to vaccination.

## Introduction

On March 11, 2020, the World Health Organization (WHO) officially declared the severe acute respiratory coronavirus 2 (SARS-CoV-2) outbreak a pandemic (1). By the end of 2020, vaccines for COVID-19 became available in the Netherlands. These vaccines showed good protection and acceptable safety profiles in clinical trials (2). However, long-term protection and occurrence of adverse events (AE) in a real-world setting need to be monitored after licensure. Furthermore, more data is required on the safety of COVID-19 vaccines in specific groups not included in clinical trials, such as people with comorbidities or pregnant women (3).

A Dutch study monitoring the first COVID-19 vaccine dose in the Netherlands, found that almost two-thirds of people experienced at least one AE after vaccination (4). AE after vaccination includes any medical occurrence and does not necessitate a causal relationship between the AE and vaccination (5). Despite this, AE following vaccination can lead to negative attitudes towards vaccination and vaccine refusal (6). Yet, it is also commonly believed that AE are positive as they are indicative of the COVID-19 vaccine eliciting a good immune response (7, 8). Evidence for this association may increase acceptance of AE after vaccination. However, the few studies assessing the relationship between COVID-19 vaccine AE and antibody concentrations have produced mixed results (8, 9).

Previous studies on the association between AE and antibody response after COVID-19 vaccination generally have small sample sizes, are performed in specific populations, e.g. healthcare professionals (HCP), and primarily include only the first and second Comirnaty vaccine doses. We use data on AE and antibody response after vaccination from a prospective cohort study performed in the Netherlands (VAccine Study COvid-19 (VASCO)) that has included a large number of individuals from the general Dutch population and has collected information on Vaxzevria, Spikevax, Comirnaty and Jcovden vaccination, as well as information on specific groups such as those with comorbidities. Therefore, this study could provide valuable evidence for the relationship between AE and antibody response.

This study aims to investigate solicited AE and AE for which medical care was sought after COVID-19 vaccination in the Netherlands and to determine the relationship between self-reported AE and anti-SARS-CoV-2 antibody concentrations prior to and after vaccination.

## Methods

### Study design

This study was performed within VASCO, an ongoing population-based prospective cohort study with the primary objective to estimate vaccine effectiveness (VE) of COVID-19 vaccines used in the Netherlands (10). Between 3 May 2021 and 15 December 2021, community-dwelling Dutch adults aged 18 to 85 years who were able to read, understand and write Dutch were included. Data was collected through digital questionnaires available to participants in a mobile app or website. At baseline, participants completed a questionnaire on sociodemographic variables, health status, COVID-19 vaccination status and SARS-CoV-2 infection test results. During follow-up, participants were asked to complete a monthly questionnaire in the first year, and three-monthly questionnaires in years two to five on health status, COVID-19 vaccination status and SARS-CoV-2 testing. Furthermore, participants were asked to notify in the app when they received a COVID-19 vaccination or tested positive for SARS-CoV-2. Participants received free self-tests during the study from April 2022 onwards when nationwide testing was scaled down. A questionnaire on solicited AE and AE for which medical care was sought was made available to participants one month after each COVID-19 vaccination.

Serum to measure anti-SARS-CoV-2 antibody concentrations was obtained by a self-collected finger-prick blood sample at baseline, six and twelve months, and one month after primary vaccination series.

The current analysis includes AE questionnaire data collected until 14 November 2022 and serology data until 6 November 2022.

### Study population

Participants with a completed AE questionnaire within two months following vaccination were included. Additional in- and exclusion criteria were applied according to each study objective. To investigate the relationship between AE and pre-vaccination anti-SARS-CoV-2 antibody concentrations a serology sample within eight weeks before vaccination was required, and participants were excluded if they reported a positive SARS-CoV-2 test between blood sampling and vaccination. To investigate the relationship between AE and post-vaccination anti-SARS-CoV-2 antibody concentrations a serology sample within eight weeks after vaccination was required, and participants were excluded if they had a history of SARS-CoV-2 infection based on a self-reported positive SARS-CoV-2 test or serology results.

### Data collection

#### Adverse events

The questionnaire included questions on the occurrence of overall injection site AE (e.g. pain, redness and swelling at the injection site or axillary region) and overall systemic AE (e.g. headache, myalgia, fatigue, nausea, diarrhoea and other symptoms of malaise), duration of AE (<-1 day, 1-2 days, 3-4 days, 5 or more days), and severity of AE (10-point Likert scale). We also asked whether participants sought medical attention for an AE. The latter questions for which medical care was sought were included in the questionnaire from the start of the study in May 2021, while the questions on solicited AE were added in August 2021. All adverse events were self-reported and not medically confirmed.

Three types of participants were identified: those with only injection site AE, only systemic AE or both. AE scores were determined by calculating the product of the duration and severity. For participants with both injection site and systemic AE, the sum of the AE scores was calculated. We used duration scores of 0.5, 1.5, 3.5 and 6 for the four duration categories. AE scores of each group were then categorised as mild or moderate severity. Mild severity was defined as a AE score below or equal to the median score of the group; moderate severity was defined as a AE score above the median score of the group. Categorization was performed by dose.

#### Antibody detection

Serum samples were tested using the Elecsys Anti-SARS-CoV2 S and Anti-SARS-CoV-2 assays (Roche Diagnostics, Mannheim, Germany). These are electrochemiluminescence immunoassays measuring total Ig levels against the receptor binding domain (RBD) domain of the Spike protein (S-antibodies) and the nucleoprotein (N-antibodies) of SARS-CoV-2 to distinguish between vaccine-induced antibodies (S-antibodies) and infection-induced antibodies (N-antibodies). Results are expressed in binding antibody units (BAU/ml).

### Data analysis

For solicited AE, the percentage of subjects who reported injection site AE and systemic AE were presented overall and stratified by age group (18-59 vs. 60-85), sex (male, female, other), vaccine dose (range 1-5), vaccine product (Comirnaty, JCovden, Spikevax, Vaxzevria), and medical risk group. Participants were categorized as belonging to a medical risk group if they had one or more of the following conditions: asplenia, cancer (in the past, currently treated, currently untreated), cardiovascular disease, diabetes, immune disorder, kidney disease, liver disease, lung disease or asthma, neurological disease, organ or bone marrow transplantation. The percentage of subjects who sought medical care concerning a potential AE were presented overall and stratified by age group, sex, medical risk group, vaccine dose and vaccine product. AE for which medical care was sought were categorized by the Netherlands Pharmacovigilance Centre Lareb using medical terminology dictionary for regulatory activities (MedDRA) higher level terms (HLT) and frequencies were presented. Potential determinants of AE occurrence were investigated using multivariable logistic regression analysis. Variables included in the model were age group, sex, medical risk group, vaccine product and vaccine dose. As data on JCovden and Vaxzevria vaccines was limited, this analysis was restricted to mRNA vaccines (Comirnaty and Spikevax).

We performed additional logistic regression analyses, stratified by dose, to investigate the association between pre-vaccination S-antibody concentrations (BAU/ml) and injection site AE, systemic AE and AE for which medical care was sought. Pre-vaccination S-antibody concentrations (BAU/ml) were categorized into quartiles, separately for each dose. We adjusted for time between blood sampling and vaccination (partly adjusted model) and additionally for vaccine product, sex and age group (fully adjusted model). Analysis was performed on participants having received an mRNA vaccine as dose two, three or four as there was limited data on dose one as AE questions were included since August 2021.

Multivariable linear regression analysis was performed, stratified by dose, to assess the relationship between AE and post-vaccination log-transformed S-antibody concentrations. Covariates in each model were age group, sex, medical risk group, BMI, vaccine product, and time between blood sampling and vaccination. Three models were built with log-transformed S-antibody concentrations as dependent variable and the following independent variables: 1) the occurrence of any AE (basic model), 2) the type of AE (injection site AE, systemic AE or both) (extended model), and 3) the severity of AE (mild or moderate) per AE type (combined model). Analysis was restricted to participants having received a mRNA vaccine as dose two or three, and additionally participants of 60 years or older having received a mRNA vaccine as dose four. Data was limited for the younger age group (18-59) receiving dose four as for this dose the vaccination policy in the Netherlands was to target people over 70 years of age, immunocompromised persons and adults with Down syndrome (11). We present geometric mean concentration (GMC) ratios with 95% confidence intervals (CI).

Statistical analyses were performed in RStudio (v4.2.2) using the tidyverse (v1.3.2) and geepack (v1.3.9) packages (12-14).

### Ethics

The VASCO study protocol was approved by the Medical Ethics Committee of Stichting Beoordeling Ethiek Biomedisch Onderzoek (BEBO), Assen, the Netherlands. The VASCO study is conducted according to the principles of the Declaration of Helsinki. Written informed consent was obtained from all participants.

## Results

### Study population

A total of 47,947 unique AE questionnaires were completed by 28,032 participants between 10 May 2021 and 14 November 2022, of which 31,775 questionnaires were completed between 24 August 2021 and 14 November 2022, and therefore included data on injection site and systemic AE (Table 1). For 91.9% of vaccines administered during the study an AE questionnaire was completed.

**Table 1.** Number of questionnaires and participants included for each research objective.

|  | (Determinants of) (un)solicited AE |  | Pre-vaccination serology |  | Post-vaccination serology |  |
| --- | --- | --- | --- | --- | --- | --- |
|  | Questionnaires (%) | Participants | Questionnaires (%) | Participants | Questionnaires (%) | Participants |
|  | n = 47,947 | n = 28,032 | n = 10,918 | n = 10,183 | n = 2,543 | n = 2,515 |
| **Age group** |  |  |  |  |  |  |
| 18-59 | 23,082 (48.1%) | 14,084 | 6,005 (55%) | 5,719 | 1,206 (47.4%) | 1,197 |
| 60-85 | 24,830 (51.8%) | 13,920 | 4,910 (45%) | 4,461 | 1,336 (52.5%) | 1,317 |
| Missing | 35 (0.1%) | 28 | 3 (0.0%) | 3 | 1 (0.0%) | 1 |
| **Sex** |  |  |  |  |  |  |
| Male | 17,047 (35.6%) | 10,060 | 3,844 (35.2%) | 3,599 | 844 (33.2%) | 836 |
| Female | 30,869 (64.4%) | 17,955 | 7,074 (64.8%) | 6,584 | 1,699 (66.8%) | 1,679 |
| Other | 31 (0.1%) | 17 | NA | NA | NA | NA |
| **Medical risk group<sup>1</sup>** |  |  |  |  |  |  |
| No | 32,997 (68.8%) | 19,670 | 7,826 (71.7%) | 7,317 | 1,827 (71.8%) | 1,809 |
| Yes | 14,950 (31.2%) | 8,362 | 3,092 (28.3%) | 2,866 | 716 (28.2%) | 706 |
| **Vaccine product** |  |  |  |  |  |  |
| Comirnaty | 22,698 (47.3%) | 16,385 | 4,880 (44.7%) | 4,735 | 1,381 (54.3%) | 1,371 |
| JCovden | 848 (1.8%) | 846 | NA | NA | NA | NA |
| Spikevax | 19,147 (39.9%) | 14,482 | 5,950 (54.5%) | 5,740 | 1,144 (45.0%) | 1,139 |
| Vaxzevria | 3,231 (6.7%) | 2,582 | NA | NA | NA | NA |
| Missing | 2,023 (4.2%) | 1,973 | 88 (0.8%) | 86 | 18 (0.7%) | 18 |
| **Vaccine dose** |  |  |  |  |  |  |
| 1 | 9,121 (19.0%) | 9,121 | NA | NA | NA | NA |
| 2 | 10,963 (22.9%) | 10,963 | 2,518 (23.1%) | 2,518 | 701 (27.6%) | 701 |
| 3 | 15,601 (32.5%) | 15,601 | 6,984 (64.0%) | 6,981 | 1,126 (44.3%) | 1,126 |
| 4 | 8,779 (18.3%) | 8,779 | 1,416 (13.0%) | 1,416 | 716 (28.2%) | 716 |
| 5 | 2,219 (4.6%) | 2,219 | NA | NA | NA | NA |
| Missing | 1,264 (2.6%) | 1,264 | 0 (0.0%) | 0 | 0 (0.0%) | 0 |

<sup>1</sup> Medical risk group: asplenia; cancer (in the past, currently treated, currently untreated); cardiovascular disease; diabetes; immune disorder; kidney disease; liver disease; lung disease or asthma; neurological disease; organ or bone marrow transplantation.

The association between pre-vaccination S-antibody levels and AE occurrence was assessed using 10,918 questionnaires, completed by 10,183 participants, for which a blood sample within eight weeks before vaccination was available. Eleven participants were excluded because they reported infection between blood sampling and subsequent vaccination. The relationship between AE and post-vaccination S-antibody response was assessed using 2,543 AE questionnaires, completed by 2,515 SARS-CoV-2 naive participants, for which a blood sample between 3-8 weeks after vaccination was available. Overall, more women than men were included and the majority of participants received Comirnaty or Spikevax.

### Solicited AE and AE for which medical care was sought

Injection site AE were reported in 42.4% of questionnaires and systemic AE in 33.8% of questionnaires (Table 2). Questionnaires completed by participants aged 18-59 years reported more injection site AE (49.3% vs. 36.9%) and systemic AE (41.8% vs. 27.3%) than questionnaires by participants aged 60-85 years. Questionnaires completed by women reported more injection site AE (50% vs. 28.8%) and systemic AE (38.4% vs. 25.5%) than questionnaires by men. Injection site AE were reported most often in questionnaires completed after vaccination with Comirnaty (43.7%) and systemic AE after vaccination with Jcovden (42.9%). Injection site AE ranged between 52.9% after the first dose and 32.8% after the fifth dose. The percentage of participants experiencing systemic AE decreased after each dose from 46.3% after the first dose to 25.6% after the fifth dose. However, this trend disappeared when data were stratified by age group (Supplementary Figure 1 and 2). The median duration of injection site AE was two days and median severity was three on a 10-point Likert scale. For systemic AE, the median duration was two days and median severity was four on a 10-point Likert scale.

**Table 2.** Reported injection site AE, systemic AE and AE for which medical care was sought by participant characteristics.

|  | Injection site AE<br>(n questionnaires =<br>31,775) | Systemic AE<br>(n questionnaires<br>= 31,775) | AE for which<br>medical care was<br>sought<br>(n questionnaires =<br>47,947) |
| --- | --- | --- | --- |
| **Reported AE** | n = 13,479<br>(42.4%) | n = 10,725<br>(33.8%) | n = 1,034<br>(2.2%) |
| **Sex** |  |  |  |
| Men | 3,275 (28.8%) | 2,893 (25.5%) | 252 (1.5%) |
| Women | 10,197 (50.0%) | 7,824 (38.4%) | 783 (2.5%) |
| Other | 7 (38.9%) | 8 (44.4%) | 2 (6.5%) |
| **Age group** |  |  |  |
| 18-59 | 6,950 (49.3%) | 5,897 (41.8%) | 586 (2.5%) |
| 60-85 | 6,521 (36.9%) | 4,826 (27.3%) | 446 (1.8%) |
| Missing | 8 (44.4%) | 2 (11.1%) | 2 (5.7%) |
| **Medical risk group** |  |  |  |
| No | 9,275 (42.6%) | 7,520 (34.6%) | 611 (1.9%) |
| Yes | 4,204 (41.9%) | 3,205 (32.0%) | 423 (2.8%) |
| **Vaccine product** |  |  |  |
| Comirnaty | 5,499 (43.7%) | 4,448 (35.3%) | 494 (2.2%) |
| Jcovden | 21 (30.0%) | 30 (42.9%) | 23 (2.7%) |
| Spikevax | 7,494 (42.6%) | 5,825 (33.1%) | 336 (1.8%) |
| Vaxzevria | 51 (25.6%) | 37 (18.6%) | 137 (4.2%) |
| Missing | 414 (31.3%) | 385 (29.1%) | 44 (2.2%) |
| **Vaccine dose** |  |  |  |
| 1 | 137 (52.9%) | 120 (46.3%) | 318 (3.5%) |
| 2 | 1,879 (45.8%) | 1,711 (41.7%) | 266 (2.4%) |
| 3 | 7,324 (47.0%) | 5,682 (36.5%) | 287 (1.8%) |
| 4 | 3,144 (35.8%) | 2,384 (27.2%) | 100 (1.1%) |
| 5 | 727 (32.8%) | 569 (25.6%) | 32 (1.4%) |
| Missing | 268 (31.7%) | 259 (30.6%) | 31 (2.5%) |

In 1,034 questionnaires (2.2%), participants reported seeking medical care for a potential AE, of which the majority were women (n = 783, 76%) and aged 18-59 years (n = 586, 57%). AE for which medical care was sought were reported most often after Vaxzevria (4.2%) and after the first dose (3.5%) (Table 2).

Logistic regression showed that younger age (18-59 years), female sex and belonging to a medical risk group were associated with higher frequency of injection site AE, systemic AE and AE for which medical care was sought reporting (Table 3). Additionally, receiving Spikevax was associated with higher frequency of injection site AE (OR: 1.30, 95%CI: 1.23; 1.37) and systemic AE (OR: 1.28, 95%CI: 1.21; 1.35) compared to Comirnaty. The odds of reporting injection site AE, systemic AE or AE for which medical care was sought decreased between the first and fifth doses.

**Table 3.**
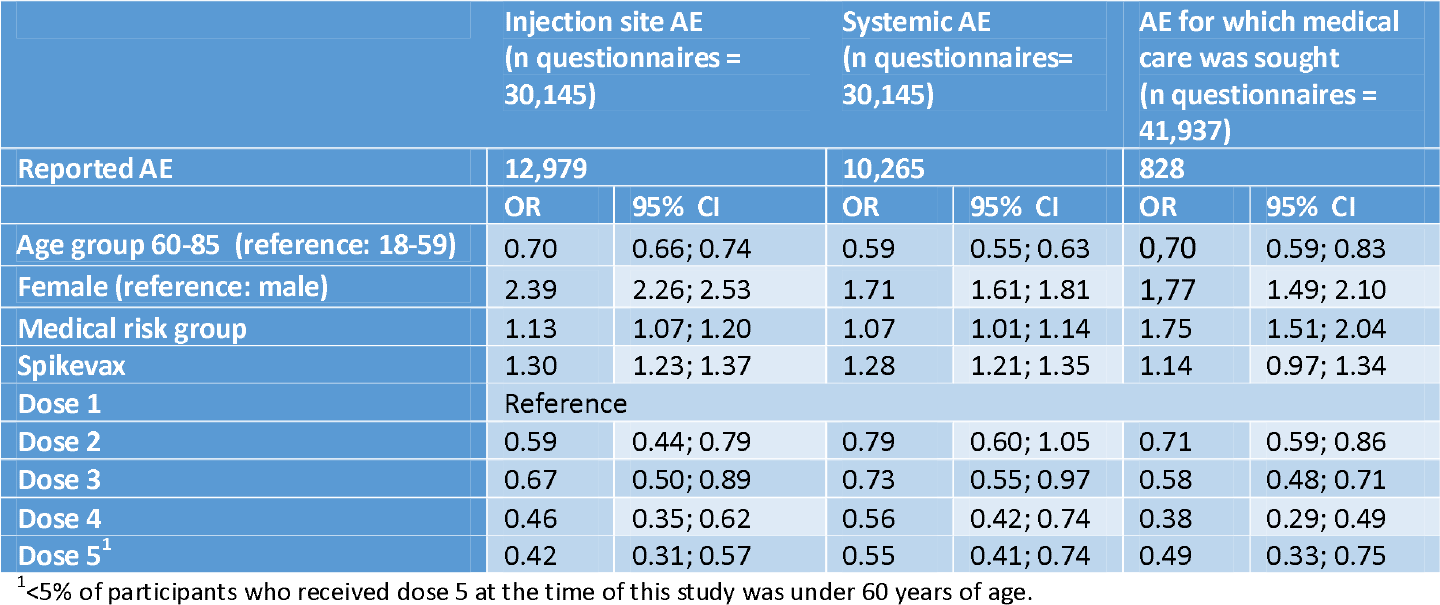
Multivariable logistic regression analysis estimating determinants for occurrence of AE. All variables were included in the model.

Participants with certain underlying conditions or medication use more often reported AE compared to those without and the frequency and type of reported AE among them also differed by age group (Supplementary Figures 3 and 4). More specifically, in both age groups, participants with gastrointestinal disease or lung disease or asthma, and participants who used gastroprotective medication reported more injection site and systemic AE than those without underlying conditions or medication use. Participants in both age groups with gastrointestinal disease or lung disease or asthma and participants who used immunosuppressive medication reported more AE for which medical care was sought (Supplementary Figures 3 and 4).

The three most frequently reported AE for which medical care was sought were headache, fatigue and fever (Supplementary Figure 5).

### Pre-vaccination S-antibody concentrations and AE

Logistic regression showed that the odds for experiencing systemic AE after dose 2 was significantly increased for those with pre-vaccination S-antibody concentrations in the third (fully adjusted OR: 1.41, 95%CI: 1.06; 1.88) and fourth (OR: 1.95, 95% CI: 1.46; 2.60) quartiles compared to those in the lowest quartile (Table 4). The odds of experiencing systemic AE after dose 3 increased with increasing pre-vaccination S-antibody concentrations: the fully adjusted OR increased from 1.26 (95%CI: 1.08;1.45) in the second quartile to 1.98 (95%CI: 1.70;2.31) in the fourth quartile. For systemic AE after dose 4, no association was observed with pre-vaccination concentrations. Some statistically significant associations were observed for pre-vaccination S-antibody concentrations and injection site AE and AE for which medical care was sought in the partly adjusted model, but results were not statistically significant when fully adjusted for time between blood sampling and vaccination, vaccine product, sex and age.

**Table 4.**
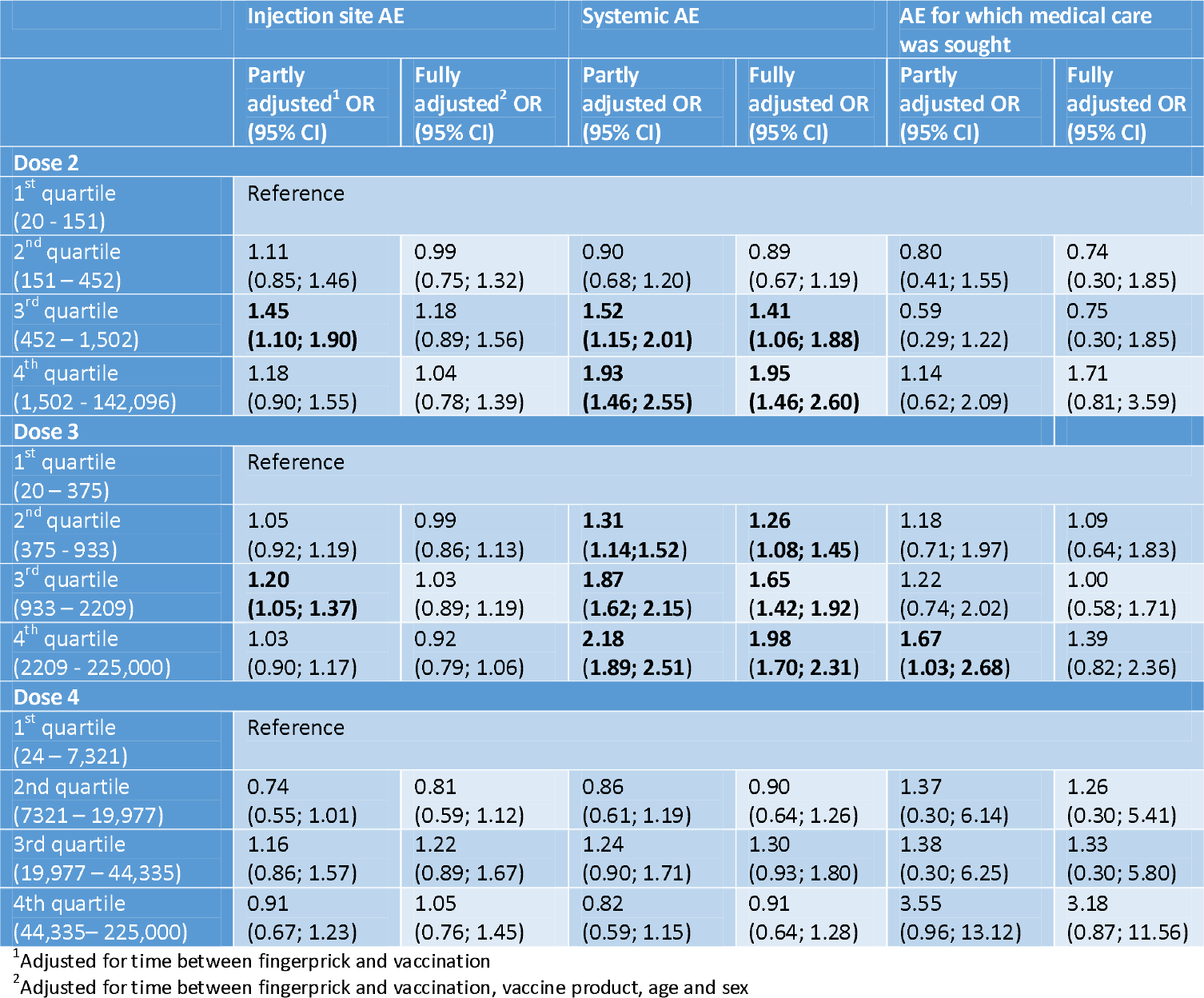
Partly and fully adjusted odds ratios with 95% confidence intervals for the association between pre-vaccination S-antibody concentrations (BAU/ml) in quartiles and type of AE stratified by dose.

|  | Injection site AE |  | Systemic AE |  | AE for which medical care was sought |  |
| --- | --- | --- | --- | --- | --- | --- |
|  | Partly adjusted<sup>1</sup> OR (95% CI) | Fully adjusted<sup>2</sup> OR (95% CI) | Partly adjusted OR (95% CI) | Fully adjusted OR (95% CI) | Partly adjusted OR (95% CI) | Fully adjusted OR (95% CI) |
| **Dose 2** |  |  |  |  |  |  |
| 1<sup>st</sup> quartile (20 - 151) | Reference |  |  |  |  |  |
| 2<sup>nd</sup> quartile (151 – 452) | 1.11 (0.85; 1.46) | 0.99 (0.75; 1.32) | 0.90 (0.68; 1.20) | 0.89 (0.67; 1.19) | 0.80 (0.41; 1.55) | 0.74 (0.30; 1.85) |
| 3<sup>rd</sup> quartile (452 – 1,502) | **1.45 (1.10; 1.90)** | 1.18 (0.89; 1.56) | **1.52 (1.15; 2.01)** | **1.41 (1.06; 1.88)** | 0.59 (0.29; 1.22) | 0.75 (0.30; 1.85) |
| 4<sup>th</sup> quartile (1,502 - 142,096) | 1.18 (0.90; 1.55) | 1.04 (0.78; 1.39) | **1.93 (1.46; 2.55)** | **1.95 (1.46; 2.60)** | 1.14 (0.62; 2.09) | 1.71 (0.81; 3.59) |
| **Dose 3** |  |  |  |  |  |  |
| 1<sup>st</sup> quartile (20 – 375) | Reference |  |  |  |  |  |
| 2<sup>nd</sup> quartile (375 - 933) | 1.05 (0.92; 1.19) | 0.99 (0.86; 1.13) | **1.31 (1.14; 1.52)** | **1.26 (1.08; 1.45)** | 1.18 (0.71; 1.97) | 1.09 (0.64; 1.83) |
| 3<sup>rd</sup> quartile (933 – 2209) | **1.20 (1.05; 1.37)** | 1.03 (0.89; 1.19) | **1.87 (1.62; 2.15)** | **1.65 (1.42; 1.92)** | 1.22 (0.74; 2.02) | 1.00 (0.58; 1.71) |
| 4<sup>th</sup> quartile (2209 - 225,000) | 1.03 (0.90; 1.17) | 0.92 (0.79; 1.06) | **2.18 (1.89; 2.51)** | **1.98 (1.70; 2.31)** | **1.67 (1.03; 2.68)** | 1.39 (0.82; 2.36) |
| **Dose 4** |  |  |  |  |  |  |
| 1<sup>st</sup> quartile (24 – 7,321) | Reference |  |  |  |  |  |
| 2nd quartile (7321 – 19,977) | 0.74 (0.55; 1.01) | 0.81 (0.59; 1.12) | 0.86 (0.61; 1.19) | 0.90 (0.64; 1.26) | 1.37 (0.30; 6.14) | 1.26 (0.30; 5.41) |
| 3rd quartile (19,977 – 44,335) | 1.16 (0.86; 1.57) | 1.22 (0.89; 1.67) | 1.24 (0.90; 1.71) | 1.30 (0.93; 1.80) | 1.38 (0.30; 6.25) | 1.33 (0.30; 5.80) |
| 4th quartile (44,335 – 225,000) | 0.91 (0.67; 1.23) | 1.05 (0.76; 1.45) | 0.82 (0.59; 1.15) | 0.91 (0.64; 1.28) | 3.55 (0.96; 13.12) | 3.18 (0.87; 11.56) |

<sup>1</sup>Adjusted for time between fingerprick and vaccination
<sup>2</sup>Adjusted for time between fingerprick and vaccination, vaccine product, age and sex

### Adverse events and post-vaccination S-antibody concentrations

GMCs post-vaccination were higher in participants with AE compared to those without AE (5,438 vs. 4,022 BAU/ml after dose 2; 20,510 vs. 15,553 BAU/ml after dose 3; 23,581 vs. 20,948 BAU/ml after dose 4).

After dose 2, a statistically significant GMC ratio of 1.24 (95%CI: 1.02-1.51; figure 1) was observed for occurrence of mild injection site and systemic AE (combined model) compared to no AE. There were no other significant associations.

**Figure 1.**
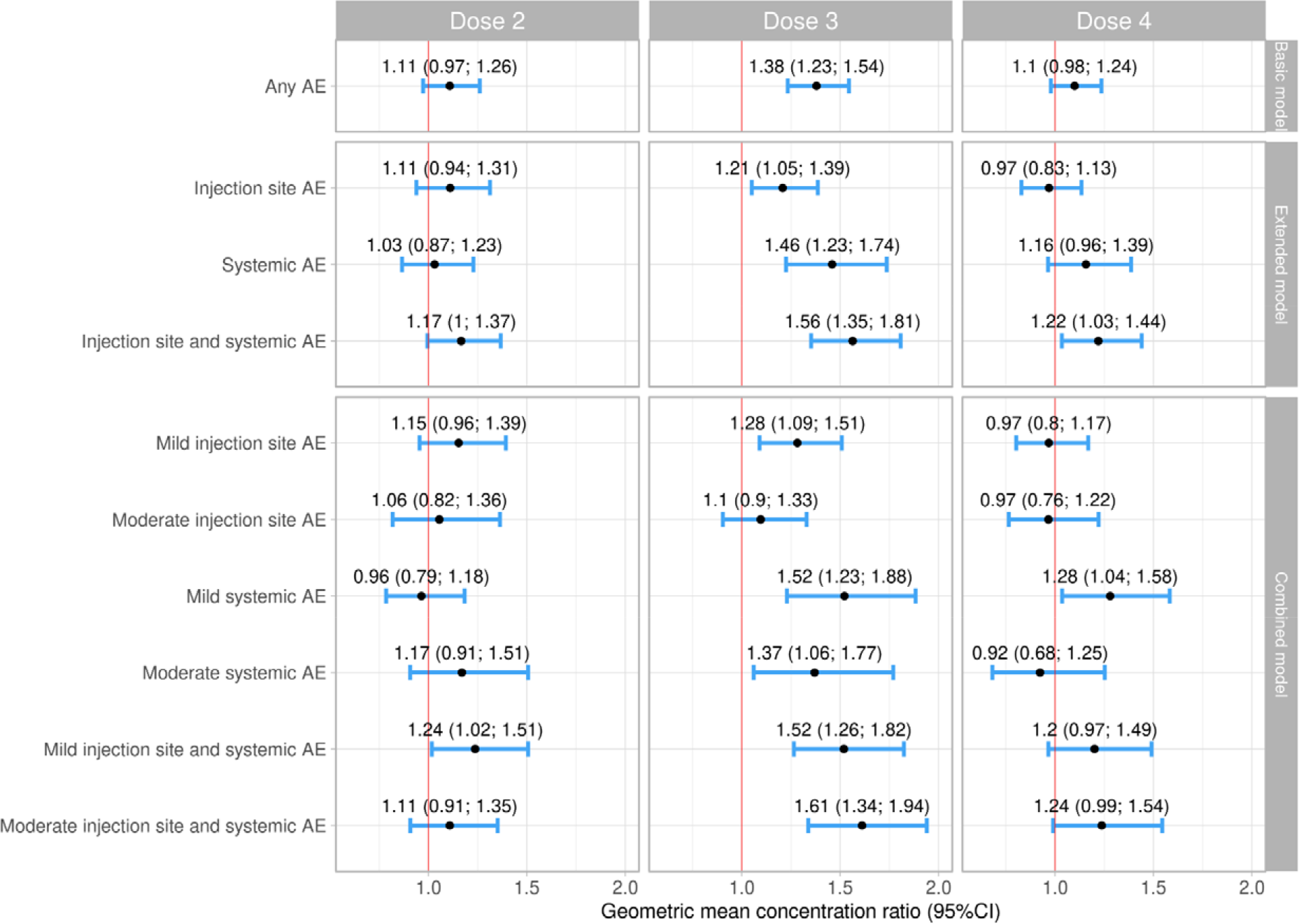
Geometric mean concentration ratios with 95% confidence intervals of post-vaccination S-antibody concentrations for any AE, type of AE, and type and severity of AE.

Occurrence of any AE after dose 3 (basic model) was associated with significantly higher antibody concentrations post-vaccination (GMC ratio: 1.38, 95%CI 1.23-1.54). Compared to experiencing no AE, the highest GMC ratio was observed for those experiencing both injection site and systemic AE (GMC ratio: 1.56, 95%CI: 1.35-1.81). In the combined model, estimated differences in S-antibody concentrations were highest for those experiencing moderate injection site and systemic AE (GMC ratio: 1.61, 95%CI: 1.34-1.94) compared to no AE.

After dose 4, experiencing both injection site and systemic AE (GMC ratio: 1.22, 95% CI: 1.03-1.44) and mild systemic AE (GMC ratio: 1.28, 95%CI: 1.04-1.58) were associated with higher S-antibody concentrations.

## Discussion

In this study, injection site AE were reported in approximately 42% of the questionnaires completed after COVID-19 vaccination, and systemic AE in almost 34% of the questionnaires. The majority of reported AE was rated less than a four on a 10-point Likert scale and resolved within three days. Female sex, individuals aged 18-59 years compared to older individuals, individuals belonging to a medical risk group, and those vaccinated with Spikevax vaccine (vs. Comirnaty) more often reported injection site AE as well as systemic AE. AE for which medical attention was sought were reported in approximately 2% of questionnaires completed after COVID-19 vaccination.

We found that higher pre-existing serum S-antibody concentrations were associated with a higher risk of AE. Specifically, after the third dose and partly after the second dose, we found an association between higher pre-vaccination antibody levels and systemic AE, but not with injection site or AE for which medical care was sought.

We also found that after the third mRNA dose post-vaccination S-antibody concentrations were up to 1.6 times higher in participants who experienced any AE compared to those who experienced no AE. After the second dose, we only found an association for those experiencing mild injection site and systemic AE and after the fourth dose in people over 60 years, only for those with both injection site AE and systemic AE or mild systemic AE.

Our results on frequency, severity and determinants of injection site and systemic AE are substantiated by other studies (4, 15). Rolfes et al (2021) performed a cohort event monitoring study in the Netherlands and found that compared to Comirnaty, the JCovden, Vaxzevria, and Spikevax vaccines were associated with increased reporting of reactogenicity. In line with our findings, they also found increased reporting of AE in women and younger participants. Differences in AE reporting between specific groups may be due to differences in the immune response according to age and sex (6). The immune system declines with age in the process of immune senescence; vaccination elicits a lesser immune response and therefore possibly less AE in an older age group (6). Furthermore, the immunosuppressive properties of testosterone compared to estrogen influence COVID-19 vaccination response and therefore reactogenicity according to sex (6). Rolfes et al (2021) also found increased reporting of AE for several medical conditions, like we did. For some conditions, such as lung disease or asthma and cancer, this increased reporting might be explained by a higher risk for symptoms such as fever in general (4). Increased reporting might also result from more frequent contact with HCP in general or increased alertness for medical occurrences compared to the population without medical conditions. Importantly, as those with underlying conditions are at higher risk for severe COVID-19 disease, vaccination is crucial. Therefore, maintaining vaccine acceptance is especially important in this group; their ongoing contact with the health care system should be used to disseminate accurate information on AE and emphasize the importance of vaccination (16).

Our findings on AE for which medical care was sought are in correspondence with previous findings by a prospective cohort study performed in Australia (17). In our study, headache, fatigue and fever were most commonly reported AE. These AE are listed in the Summary of Product Characteristics (SmPC) of the Comirnaty and Spikevax vaccines (18, 19). These are generally mild and expected AE, but our study found that they may cause vaccinees to seek medical attention, possibly in relation to younger age and underlying conditions or medication use.

Notably, studies report an increase in AE after the second and third dose of COVID-19 vaccination as compared to the first dose (15, 20). We had limited data on injection site and systemic AE after the first dose and therefore we could not validly study this. However, compared to the first dose, systemic AE seemed actually to decrease after subsequent doses in our study. Increased AE after the second and booster doses might be due to pre-existing immunity resulting from previous vaccine doses (6). Pre-vaccination antibody levels are influenced by the interval between vaccines or infection and vaccination, with longer dosing intervals possibly leading to higher antibody responses (21). As dosing intervals differed between countries, this might also explain why some studies reported an increase in AE after the second and third dose. In our study, there was an association between pre-vaccination antibody concentrations and systemic AE after the second and third vaccine doses. Previous research on the relationship between antibody levels just before vaccination and AE has conflicting results (6). More research is needed to uncover this mechanism.

Regarding the relationship between AE and antibody response, some studies with comparable methods did find an association between second dose mRNA vaccine AE and post-vaccination S-antibody concentrations (8, 22, 23). Other research, however, is in alignment with our findings and did not find a correlation or no strong enough evidence to conclude that AE occurrence reflects higher antibody response after the second dose (24, 25). There is limited research on AE after booster COVID-19 vaccination and the relationship with antibody levels. A study from September 2022 on the booster Comirnaty vaccine reported a trend for higher antibodies in the month after vaccination in participants who experienced fever but this association was not significant (26). Although no signification association was found here, the mechanisms in which vaccination induces the immune response are also known to be involved with systemic side-effects, such as fever (6). Therefore, an association between AE and antibody response may be expected. A study on the persistence of SARS-CoV-2 antibodies related to symptoms found that those with COVID-19 like symptoms had higher antibody concentrations for IgG and IgM compared to those who were asymptomatic or had only mild symptoms, possibly due to a stronger inflammatory response (27).

In our study, we mainly found associations between AE and antibody response after the third mRNA vaccine dose. Previous research has shown increasing post-vaccination anti-S antibody concentrations for subsequent vaccine doses, with maximal immunogenicity reached after the third dose (28, 29). Two clinical trials investigating the immunogenicity of fourth mRNA vaccine doses found that participants with high levels of response before the fourth dose only had limited boosting after the fourth vaccination due to a ceiling effect (29, 30). This may explain why we saw respectively no association or weaker associations between AE and pre- and post-vaccination antibody levels after the fourth dose.

Variation between doses may also originate from adaptations made to COVID-19 vaccines due to ongoing vaccine development. Additionally, the Spikevax booster dose was halved, possibly leading to different antibody trajectories and AE occurrence compared to the full dose administered in the primary series (31).

Between studies, the mixed results on the relationship between AE and antibody response may result from differences in methodology. For example, we collected data on injection site and systemic AE overall, but S-antibody acquisition may instead be reflected by specific injection site or systemic AE. Kobashi and colleagues (2022) found a correlation with muscle and joint pain, and fever after a second dose mRNA vaccination, but not with fatigue or headache (32). Disparity between studies may also originate from varying antibody trajectories resulting from differences in vaccine schedules and vaccine products (21). Time point of blood sample measurement also differed across studies. Oyebanji et al (2021), Uwamino et al (2022) and Braun et al (2022) collected blood samples at approximately two weeks, three weeks and one month after the second dose respectively, whereas we included blood samples taken between three to eight weeks after vaccination (8, 20, 23). In addition, many studies were performed in HCP populations as opposed to the general population, but HCP were found to have higher IgG peak levels after vaccination compared to non-HCP, potentially due to the healthy worker effect (21). Moreover, a wide variety of assays was used to assess different serology outcome measures. These methodological differences decrease comparability of studies.

A strength of this study is the large study population for which extensive information was available on covariables. Also, we were able to exclude participants with a history of SARS-CoV-2 infection, which is associated with both increased reactogenicity and immunogenicity, from our analyses on the relationship between AE occurrence and antibody response (33). In addition, we included vaccinations beyond the primary vaccination schedule in our analyses, which contributes to the applicability of this study. This study also has several limitations. Firstly, AE questionnaires could be subject to recall bias, which may have caused participants to under- or overestimate their experience. However, as questionnaires were made available within a relatively short time-span of four weeks after vaccination, bias is most likely limited. Finally, while we were able to adjust for several variables, there were also possible confounders on which no information was available. For example, we were not able adjust for the (daily) use of antipyretic drugs, which has been linked to blunted SARS-CoV-2 antibody responses (6, 32). Not considering antipyretic drug use in our analyses may have overestimated the association between AE and antibody response. Additionally, the association between antibody response and AE may be confounded by pre-vaccination antibody levels. In this study, we found an association between pre-vaccination levels and the occurrence of AE after vaccination. Due to low numbers of participants with a blood sample shortly before and shortly after vaccination we could not adjust for pre-vaccination antibody levels in our analysis of the relationship between AE and post-vaccination antibody levels, which may have influenced our findings.

In conclusion, this study showed that for up to 43% of vaccine doses, AE were reported shortly after COVID-19 vaccination. Female sex, younger age, belonging to a medical risk group and receiving Spikevax were associated with experiencing AE. Higher pre-vaccination S-antibody levels were also associated with experiencing systemic AE after the second and third mRNA COVID-19 vaccine doses. In addition, experiencing AE may reflect higher antibody acquisition, as we observed after booster doses. Our research provides evidence regarding determinants for AE, including the role of pre-vaccination antibody levels, as well as evidence for the association between AE and COVID-19 vaccination antibody response.

## Data Availability

All data produced in the present study are available in aggregated and anonymized form upon reasonable request to the authors.

## Acknowledgements

We thank Christel Hoeve and Lareb for their contribution in this study. Conflict of interest: None

