## Supplemental files for "Association between common adverse events after COVID-19 vaccination and anti-SARS-CoV-2 antibody concentrations in a population-based prospective cohort study in the Netherlands"

### Appendix

#### Appendix 1

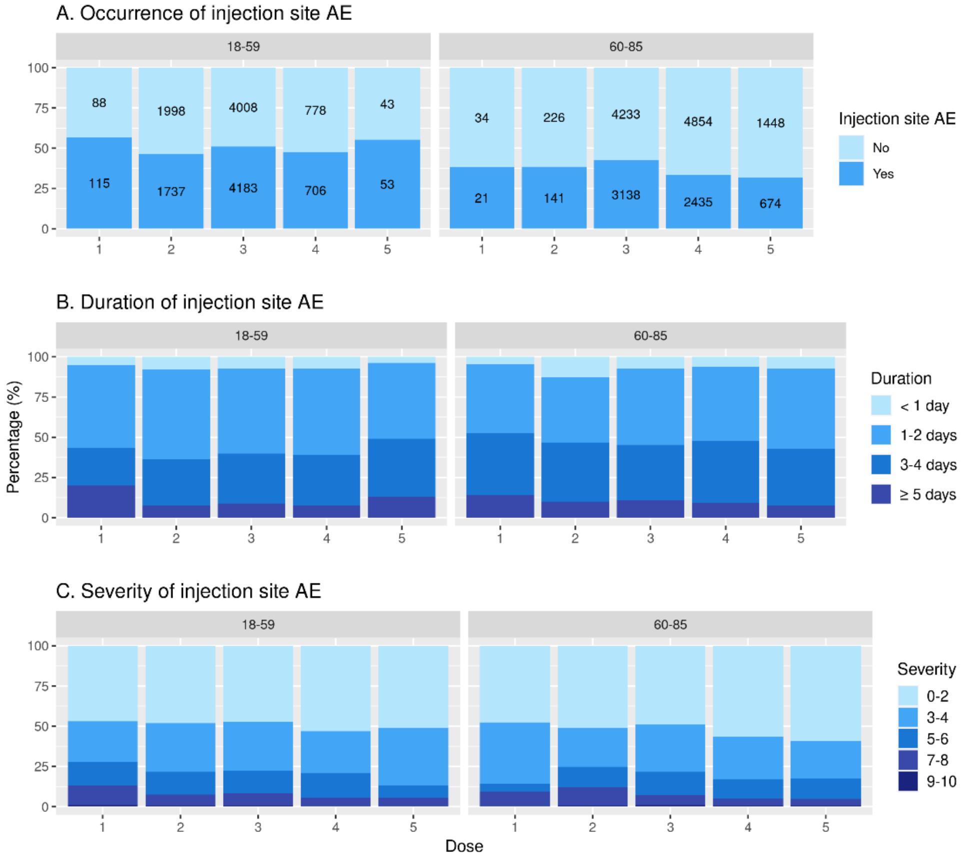


*Supplementary figure 1: Injection site AE occurrence, duration and seriousness by age group*

### Appendix 2

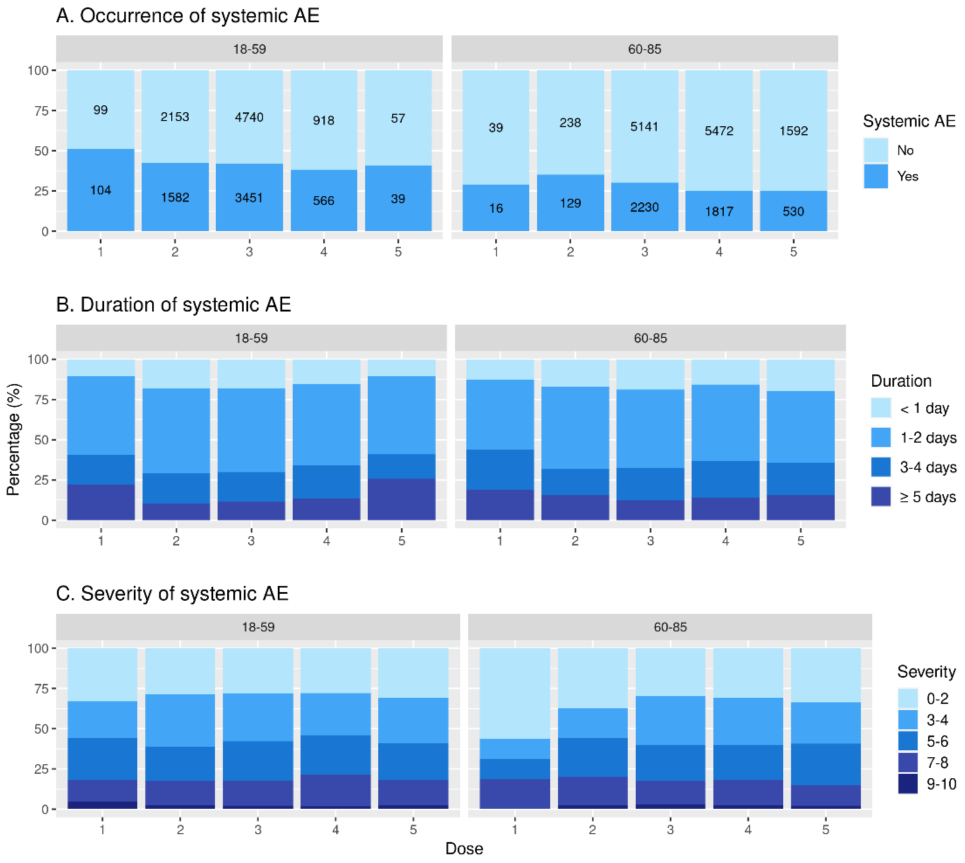


*Supplementary figure 2: Systemic AE occurrence, duration and seriousness by age group*

#### Appendix 3

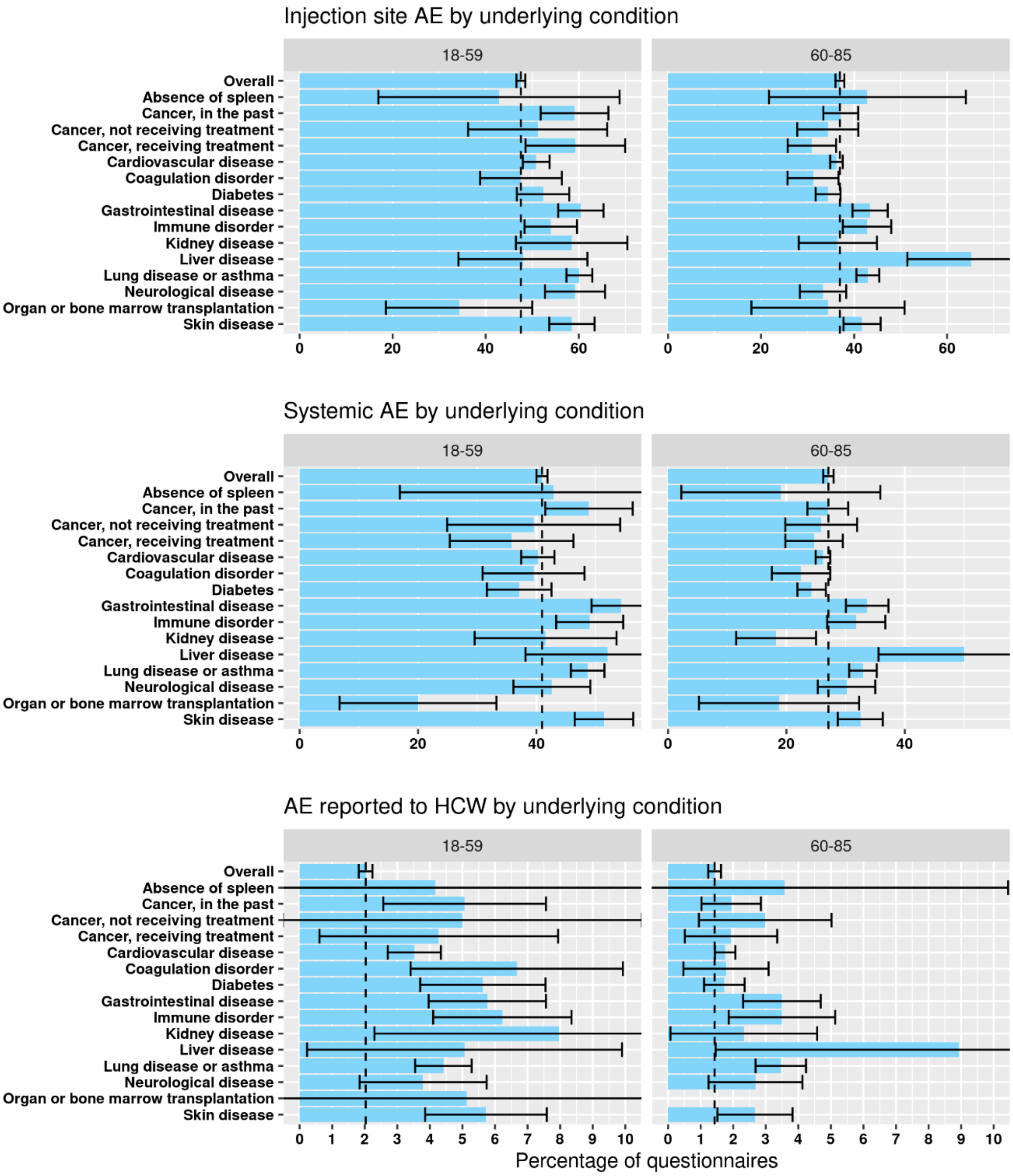


*Supplementary figure 3: Injection site AE, systemic AE and AE for which medical care was sought reporting by underlying condition*

### Appendix 4

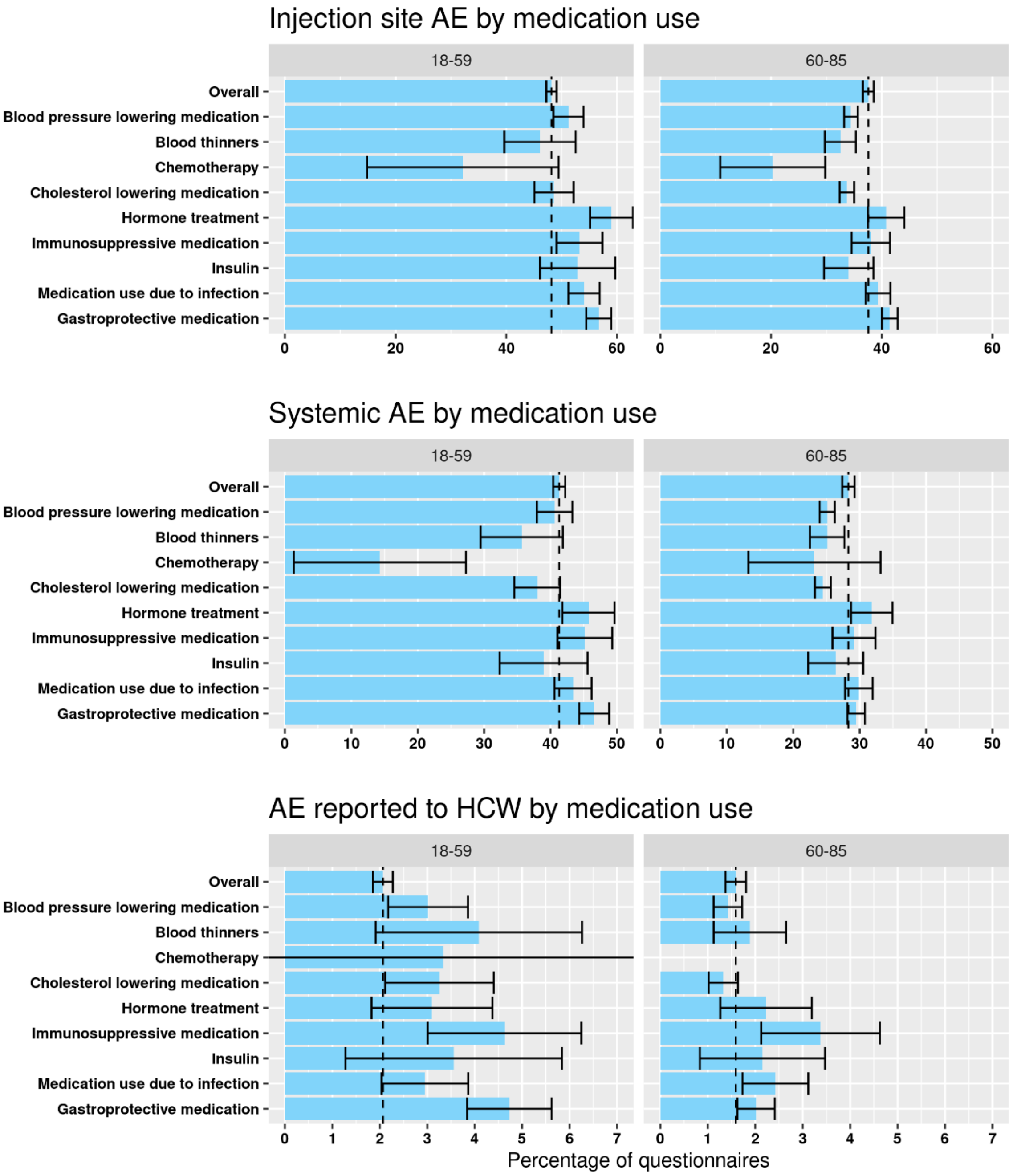


*Supplementary figure 4: Injection site AE, systemic AE and AE for which medical care was sought reporting by medication use*

#### Appendix 5

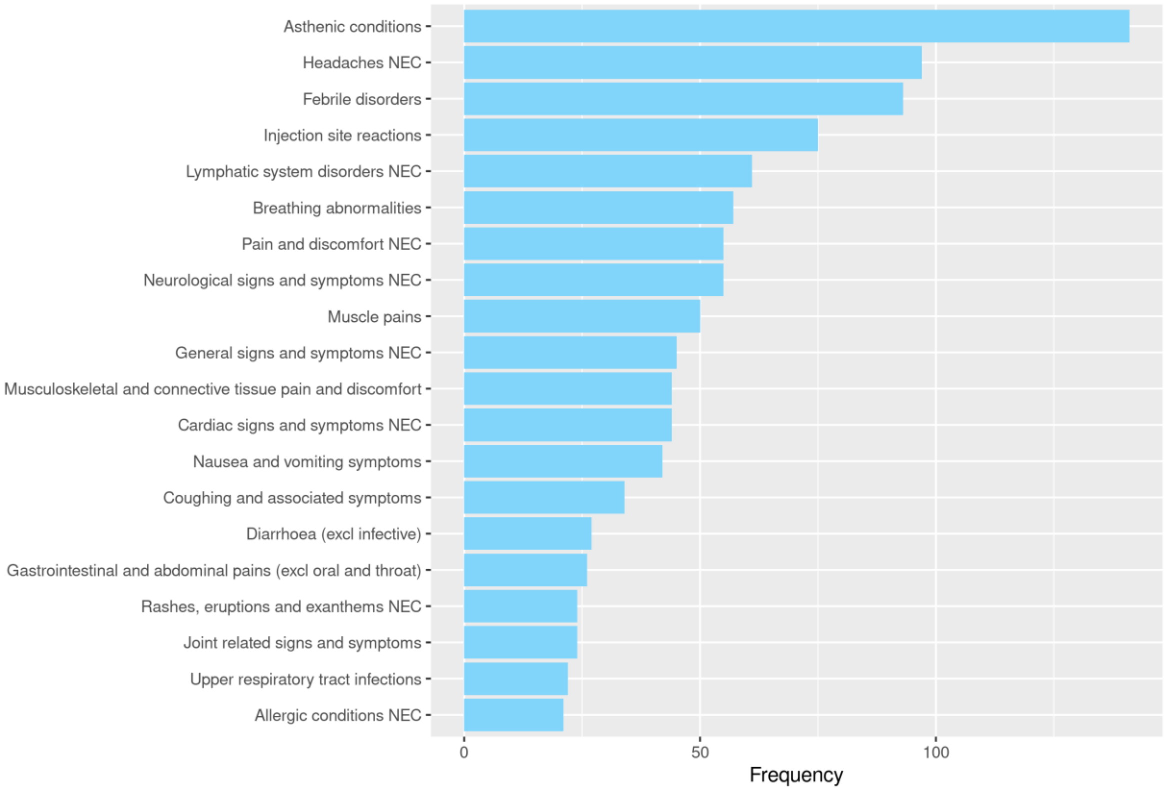


*Supplementary figure 5: Top 20 most frequently reported AE for which medical care was sought*
